# A genome-wide cross-trait analysis identifies causal relationship and shared loci of food preference with obesity

**DOI:** 10.1101/2024.06.13.24308909

**Authors:** Shi Yao, Hao Wu, Peng Bao, Long Qian, Ji-Zhou Han, Yan Wang, Si-Fan Feng, Yu-Jie Cai, Jing Guo, Xin Ke, Wei Shi, Fu-Bin Ma, Qiu-Hao Liang, Shan-Shan Dong, Yan Guo, Dong-Li Zhu, Li-Li Cui

**Affiliations:** Guangdong Key Laboratory of Age-Related Cardiac and Cerebral Diseases, Affiliated Hospital of Guangdong Medical University, Zhanjiang, Guangdong, China; Key Laboratory of Biomedical Information Engineering of Ministry of Education, Biomedical Informatics & Genomics Center, School of Life Science and Technology, Xi’an Jiaotong University, Xi’an, Shaanxi, China; Jiahui International Hospital, Shanghai, China; Department of Clinical Immunology, Xijing Hospital, Fourth Military Medical University, Xi’an, China

**Keywords:** Food preference, Obesity, Mendelian randomization, Cross-trait genetics, GWAS, Pleiotropy

## Abstract

Food preferences play a pivotal role in dietary choices and body weight regulation, yet the causal relationships and complex pathways linking food liking to obesity remain elusive. Here, we employed a two-sample Mendelian randomization (MR) analysis and pleiotropic analysis to explore the causalities and shared loci between 187 food preferences (*N*=161,625) and five obesity-related phenotypes (*N*=100,716 to 322,154). MR analysis revealed a causal association between two food-liking phenotypes and increased body mass index (BMI), specifically, onion liking (β, 0.286; 95% CI, 0.185 to 0.387; *P* = 2.80×10^−8^), and highly palatable food liking (β, 0.266; 95% CI, 0.140 to 0.391; *P* = 3.31×10^−5^). Multivariable MR analysis indicated that the impact of onion liking on BMI persisted even after conditioning on actual onion intake, suggesting a degree of independence in dietary preferences’ influence. Pleiotropic analysis under a composite null hypothesis detected 32 pleiotropic loci and six colocalized loci in these two trait pairs. Candidate pleiotropic genes associated with onion liking-BMI highlighted biological pathways primarily involved in the sensory perception of smell. These findings enhance our comprehension of the intricate relationship between food preferences and obesity.

## Introduction

Obesity has become a global epidemic, leading to a dramatic increase in the incidence of various complications ^1,2^ that adversely affect both quality of life and life expectancy^3^. The fundamental cause of obesity lies in the long-term imbalance between calories consumed and expended ^4,5^, with numerous factors involved in its development, including genetic, developmental, behavioral, and environmental influences. Many studies indicate that dietary preferences and liking food are crucial in dietary choices and body weight regulation ^6^.

Food preferences are particularly important as they strongly influence food acceptance. While human food preferences generally incline toward sweetness and away from bitterness, differences exist in the appreciation of basic taste qualities and specific foods ^6,7^. The variations in food preferences partly explain variations in short-term food intake, yet the impact of specific food liking on obesity remains debatable. For instance, highly palatable foods, often characterized by their high content of simple carbohydrates and fats, may lead individuals with a strong preference for such foods to exhibit excessive calorie consumption and weight gain due to their hedonic properties and sensory appeal ^8–12^. This hypothesis has been further supported by the positive genetic correlation between liking highly palatable foods and adverse anthropometric outcomes ^13^. However, several studies have found that obese individuals tend to prefer fats or sweetness to a similar or even lesser extent than non-obese counterparts ^14–16^. Current conclusions primarily stem from conventional observational studies, which remain challenging due to inherent limitations in observational designs, leaving the causal link between food preferences and obesity unclear. Recent large-scale genetic studies on food liking ^13^ provide crucial opportunities to investigate the causal relationship between food-liking and obesity. By leveraging the random assortment of genetic alleles as instrumental variables, Mendelian randomization (MR) can overcome confounding and reverse causation biases, providing insights into causal inference ^17^.

Weight-loss interventions focusing on reducing energy intake and increasing energy expenditure often prove ineffective in the long term ^2,18^, probably attributed to intricate hormonal, metabolic, and neurochemical adaptations that counteract weight loss efforts and facilitate weight regain. Recently, research identified that sensory food perception can rapidly promote anticipated responses to changes in energy status, which play a crucial role in controlling metabolic homeostasis ^19^. However, whether specific food preferences might impact metabolic balance without actual food consumption remains elusive. Moreover, delineating the functional genomics of cross-phenotype genetic effects can uncover fundamental aspects of pleiotropy and help identify therapeutic targets. To establish a more nuanced insight into the complex pathways linking food liking to obesity, it is crucial to identify the specific genomic variants that contribute to the shared genetic etiology of food liking and obesity.

In this study, utilizing large-scale genome-wide association study (GWAS) summary statistics, we investigated the genetic correlation and causal relationships between 187 food preferences and five obesity phenotypes, including body mass index (BMI), body fat percentage (BFP), waist-hip ratio (WHR), hip circumference (HIP), and waist circumference (WC). Subsequently, we conducted a multivariable MR analysis to estimate the direct impact of identified food preferences on obesity, conditioned on corresponding reported consumption. Additionally, we employed genome-wide pairwise trait pleiotropic analysis to sequentially explore pleiotropic associations across single-nucleotide variations, genes, and biological pathways to unravel the shared genetic basis between casual food liking and obesity. Figure 1 provides an overview of the study design. Our study aims to elucidate the mechanisms through which food liking influences obesity risk, contributing to the ongoing discourse on the multifactorial nature of obesity.

**Figure 1.**
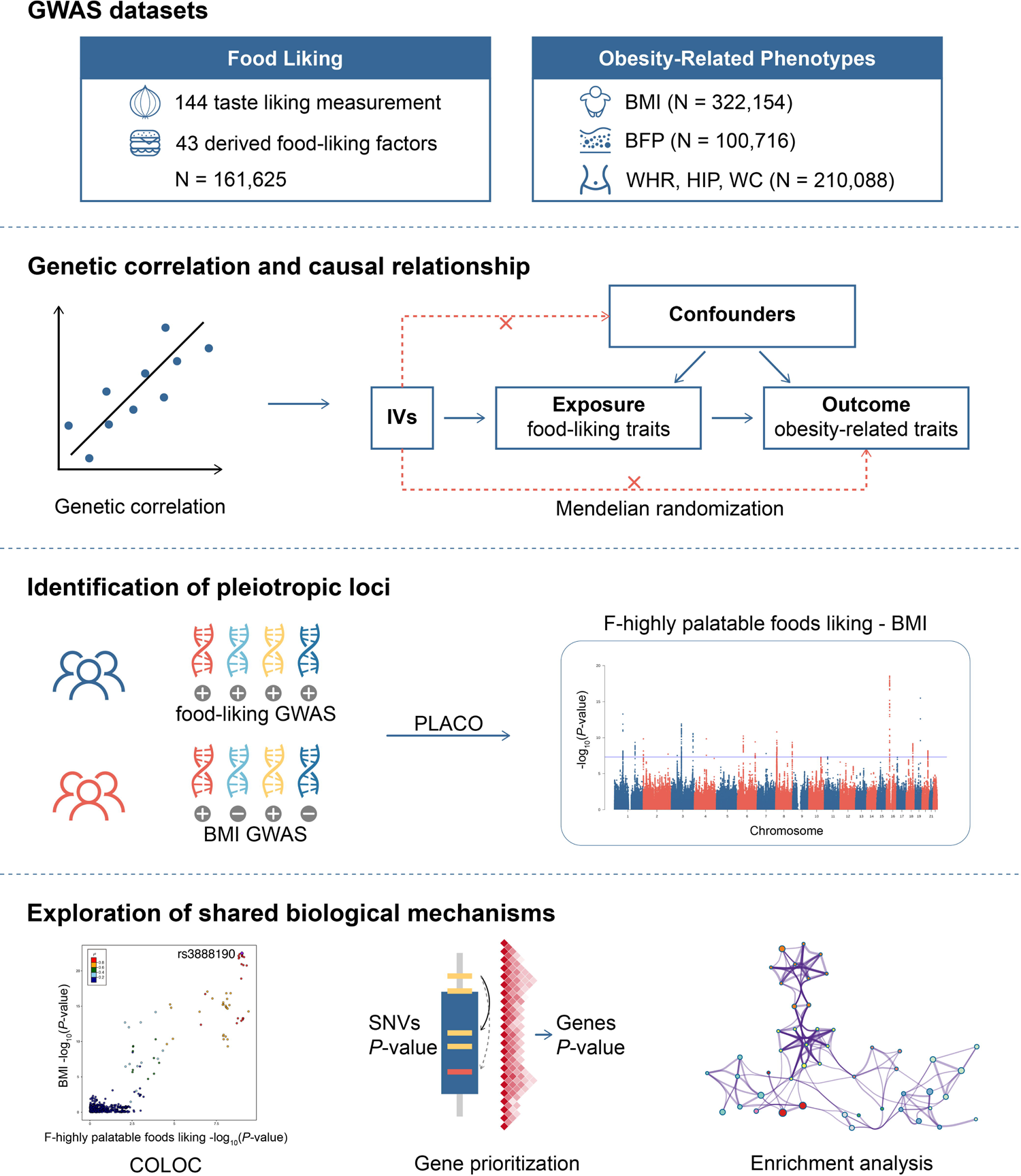
Study workflow. We conducted genetic correlation, Mendelian randomization, comprehensive pleiotropic analysis between 187 food liking traits and five obesity-related phenotypes. GWAS, genome-wide association study; BMI, body mass index; BFP, body fat percentage; WHR, waist-hip ratio; HIP, hip circumference; WC, waist circumference; IVs, instrumental variables; PLACO, pleiotropic analysis under composite null hypothesis; COLOC, colocalization.

## Methods

### GWAS data sets

Our study utilized publicly available summary-level data with ethical approval from the original studies. Detailed datasets concerning food liking and obesity-related traits are available in the original publications and summarized in Supplementary Table 1.

### Food-liking phenotypes

Genetic information regarding food-liking traits was extracted from a GWAS ^13^ conducted on 161,625 participants from the UB Biobank. This dataset included 144 detailed food and beverage preference traits, rated on a 9-point scale ranging from 1 (extremely dislike) to 9 (extremely like). Genetic correlations combined with structural equation modeling further derived 43 higher-order factor traits across three main dimensions: highly palatable, acquired, and low caloric, resulting in 187 food-liking traits.

### Obesity-related phenotypes

To mitigate weak instrument bias in MR analyses caused by participant overlap between exposure and outcome ^20^, we utilized GWAS datasets of five obesity-related phenotypes with maximized sample sizes while ensuring sample independence within the UK Biobank. Specifically, BMI statistics were obtained from a GWAS meta-analysis encompassing 322,154 individuals of European ancestry ^21^, body fat percentage from 100,716 individuals ^22^, and body fat distribution traits such as waist-hip ratio, waist circumference, and hip circumference adjusted for BMI from up to 210,088 individuals ^23^.

### Genetic correlation analyses

To enhance our understanding of the relationship between food liking and obesity, we conducted linkage disequilibrium (LD) score regression (LDSC) ^24^ to assess heritability and genome-wide genetic correlations across 935 pairs of traits before conducting MR analyses. We applied a relaxed threshold of *P* < 0.01 to encompass all traits with suggestive evidence. Given the significant number of food-liking traits and their intercorrelations, we employed PhenoSpD ^25^ to determine the independent counts of food-liking components correlated with obesity, thus addressing the issue of multiple testing. The genetic correlation matrix of food liking served as input for PhenoSpD.

### Mendelian randomization analyses

#### Instruments selection

We utilized the clump function in PLINK software ^26^ to select independent single-nucleotide polymorphisms (SNPs) from the exposure GWAS summary datasets as instrumental variables. Variants with a minor allele frequency < 0.01 were filtered out, and stringent criteria for clumping were applied with the following parameters: *r^2^* < 0.001, a window of 10,000 kb, and a significance threshold of *P* < 5×10^-8^. We utilized the European population data from the 1000 Genomes Project as the reference dataset. In cases where SNPs associated with the exposure were unavailable in the outcome datasets, proxy SNPs (*r^2^* > 0.8) were substituted. To mitigate potential confounding effects, SNPs associated with confounders such as drinking, smoking behavior, education, and socioeconomic status ^27,28^, as identified in the PhenoScanner database ^29^, were excluded from the analysis. Additionally, palindromic SNPs with intermediate allele frequencies (>0.42) were eliminated to avoid potential strand-flipping issues. Outlier pleiotropic SNPs detected by RadialMR^30^ were removed to improve the accuracy and robustness of the remaining genetic instruments. *F*-statistics were computed to assess the strength of each genetic instrument, and only SNPs with *F*-statistics > 10 were retained in the MR analysis ^31,32^. Exposures with fewer than three instrument variables were excluded from further analyses.

#### Two-sample MR analyses

We employed the inverse-variance weighted (IVW) method ^33^ as the primary approach for causal inference, aggregating Wald ratio estimates of each SNP into a single causal estimate. We reported MR estimates as log (OR) values representing the change in obesity per standard deviation increase in genetically predicted food liking. Causal analyses were simultaneously conducted using four additional methods that rely on different assumptions than IVW to address potential bias from pleiotropic instrumental variables, including robust adjusted profile score (RAPS) ^34^, weighted median ^35^, weighted mode ^36^, and MR-Egger ^37^. All the MR analyses were performed using the TwoSampleMR (version 0.4.26) R package. To enhance the reliability of the MR analyses, only estimates showing consistent direction across different methods were considered, alongside the IVW estimates retaining significance after Bonferroni correction, suggesting a potential causal association. Bonferroni correction was applied to account for multiple testing, with a significance threshold set at 5.88[×[10^−4^ (0.05 divided by 85.1, representing the independent number of food liking and obesity pairs filtered through genetic correlation analysis and instrumental variable selection). Furthermore, we extended the aforementioned MR analysis to include bidirectional causal inference to assess the causal effect of obesity on food-liking phenotypes.

#### Sensitivity Analysis

Several sensitivity analyses were performed to address potential pleiotropy issues. The intercept from MR-Egger regression was utilized to evaluate the presence of unbalanced horizontal pleiotropy, with an intercept deviation from zero indicating horizontal pleiotropy ^37^. MR-PRESSO ^38^ was employed to detect pleiotropy by comparing the observed distance of all variants to the regression line with the expected distance under the null hypothesis of no horizontal pleiotropy. Cochran’s Q statistic test ^39^, generated by different genetic variants in the fixed-effect variance weighted analysis, was used to assess heterogeneity and a *P* value of < 0.05 suggested pleiotropy. A leave-one-out analysis, which removes one variant from the analyses and re-estimates causality, was conducted to determine specific variants driving causal associations. MR Steiger tests were performed to estimate the potential reverse causal impact of obesity on food liking ^40^.

#### Multivariable MR analyses

Food-liking traits play crucial roles in dietary choices and consumption. To investigate whether specific food preferences influence metabolic balance independently of actual food consumption, we conducted a multivariable MR ^41,42^ to estimate the direct impact of food-liking traits on obesity. For food likings significantly affecting obesity and with corresponding consumption datasets, their genetic instruments were combined and clumped again to ensure SNP independence. It is important to note that we employed a relaxed *P*-value threshold of 1×10^-5^ for food intake to ensure an adequate number of instrument variables were included. SNP effects and standard errors were extracted from food-liking and consumption GWAS summary statistics and harmonized with obesity GWAS information with parameter settings consistent with univariate MR analyses.

### Identification of pleiotropic variants

The pleiotropic analysis under the composite null hypothesis (PLACO) method ^43^ was implemented to uncover potential pleiotropic effects among each of the food-liking and obesity pairs that show causal associations. PLACO provides variant-level statistical testing of pleiotropy against a composite null hypothesis, and variants with a significance threshold of *P* < 5 × 10^−8^ were considered significant pleiotropic variants. Variants with *Z^2^* > 80 for one trait were removed to avoid the influence of extremely large effect sizes. We then employed functional mapping and annotation of genetic associations (FUMA) ^44^ to determine potential pleiotropic loci. Subsequently, Bayesian colocalization analysis was conducted to explore potential shared causal variants within each pleiotropic locus ^45^. A genomic locus with a posterior probability of H4 (PP.H4) > 0.7 was defined as a colocalized locus with a potential shared causal variant, and the SNP with the largest PP.H4 within this locus was recognized as a candidate causal variant.

### Functional annotation and GSEA

#### Gene annotation

To explore the shared mechanisms of pleiotropic loci, we conducted a gene-based association analysis utilizing the multi-marker analysis of genomic annotation (MAGMA) ^46^. Specifically, MAGMA converts SNP-level associations into gene-level associations through a multiple regression method that considers LD between variants and detects multi-marker effects, with significance declared at Bonferroni-corrected *P* < 0.05. MAGMA gene-property analyses were performed across 54 tissues in GTEx v8 to identify tissue specificity regarding food liking and obesity. Genes were also annotated using FUMA ^44^, employing positional mapping, eQTL mapping, and 3D chromatin interaction mapping. Datasets for the tissues identified and available in FUMA were utilized for eQTL gene annotation and 3D chromatin interaction annotation.

#### Gene set enrichment analysis

We performed a gene set enrichment analysis using Metascape ^47^ for Gene Ontology (GO) terms and Kyoto Encyclopedia of Genes and Genomes (KEGG) pathways among genes implicated in pleiotropic loci. Metascape conducted a pathway enrichment analysis to elucidate the function of mapped genes based on MSigDB ^48^. Pathways containing at least three candidate genes, with an enrichment factor > 2 and adjusted *P* using the Benjamini-Hochberg procedure < 0.05, were considered significantly enriched pathways and grouped into clusters based on membership similarities.

## Results

### Genetic correlations between food liking and obesity

We assessed the genome-wide genetic correlations between 187 food preferences and five obesity phenotypes using summary statistics from publicly available data sources (Supplementary Table 1). Across the 935 food-liking and obesity pairs, we estimated 263 pairwise traits with nominal *P* values < 0.01 (Supplementary Table 2). As expected, we observed a positive correlation between highly palatable food liking and obesity indicators (BMI, WHR, and WC) and a negative correlation between low-caloric food liking and obesity (WC and WHR). Subsequent two-sample MR analyses were conducted to investigate the causal relationships between these pairs of traits.

### Causal effects between food liking and obesity

All single-nucleotide polymorphisms (SNPs) selected for the MR analyses are listed in Supplementary Table 3 for replication, with all *F*-statistics exceeding 30, indicating the robustness of the instrumental variables. To improve the accuracy and robustness of the genetic instruments, we excluded SNPs associated with confounding factors and outlier pleiotropic SNPs (Supplementary Tables 4-5). A total of 183 pairs of food liking and obesity, with at least three instrument variables, were included for further analyses. We employed PhenoSpD ^25^ and determined that the independent number of food liking and obesity pairs was 85.1, with a significance threshold set at 5.88[×[10^−4^ (0.05 divided by 85.1) (Supplementary Table 6).

We identified a positive causal relationship between two food-liking traits and BMI: onion liking and highly palatable food liking (Figure 2, Supplementary Table 7). Specifically, onion liking was associated with increased BMI (inverse-variance weighted (IVW) β, 0.286; 95% confidence interval (CI), 0.185 to 0.387; *P*=2.80×10^−8^), and a similar association was observed for highly palatable food liking (IVW β, 0.266; 95% CI, 0.140 to 0.391; *P*=3.31×10^−5^). Scatter plots illustrate the effects of the instruments on food liking and BMI, where colored lines represent the slopes of various regression analyses (Figure 3). MR estimates for the effects of SNPs associated with food liking on BMI are displayed in forest plots. Estimates remained consistent across MR robust adjusted profile score (RAPS), weighted median, weighted mode, and MR-Egger supporting the positive impact of liking onions and highly palatable foods on BMI (Figure 2). Sensitivity analyses revealed no evidence of pleiotropy in the causal estimates (Supplementary Tables 8-10). Analyses excluding each SNP revealed that none of the SNPs were driving the MR effects, and funnel plots indicated that potential biases were minimized with SNPs symmetrically distributed (Supplementary Figure 1). Additionally, we also identified several causal relationships at a nominally significant level, such as fatty food liking was associated with increased WC (IVW β, 0.135; 95% CI, 0.058 to 0.213; *P*=6.31×10^−4^), and strong vegetable liking was associated with decreased BMI (IVW β, −0.045; 95% CI, −0.072 to −0.018; *P*=9.64×10^−4^).

**Figure 2.**
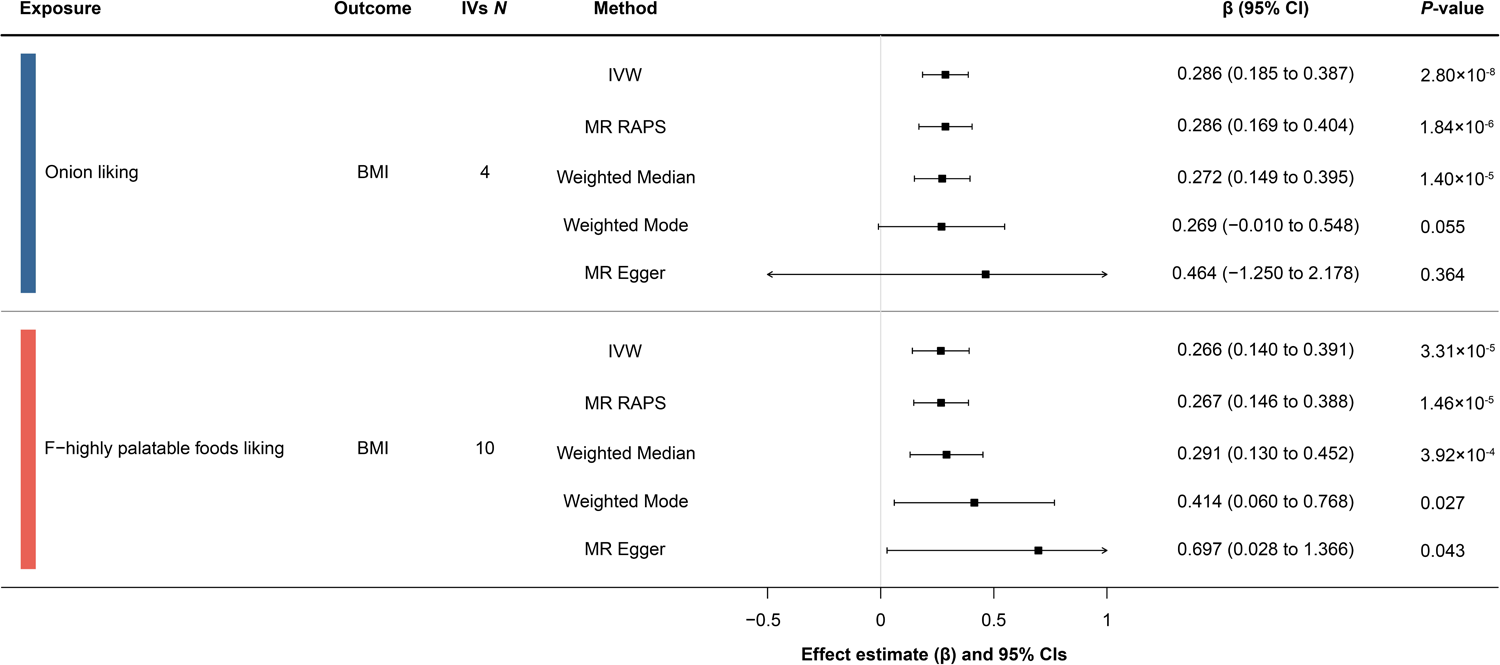
Causal effects of food liking on obesity. The forest plot of the significant causalities. The effect represents the log (OR) of obesity-related phenotype per standard deviation increase in food preference, and the error bars represent 95% CIs. Five methods were used to estimate the causal effects including inverse-variance weighted (IVW), MR robust adjusted profile score (RAPS), weighted median, weighted mode, and MR-Egger. IVW *P*-value < 5.88 × 10^-4^ after Bonferroni corrections was considered significant. IVs, instrumental variables; BMI, body mass index.

**Figure 3.**
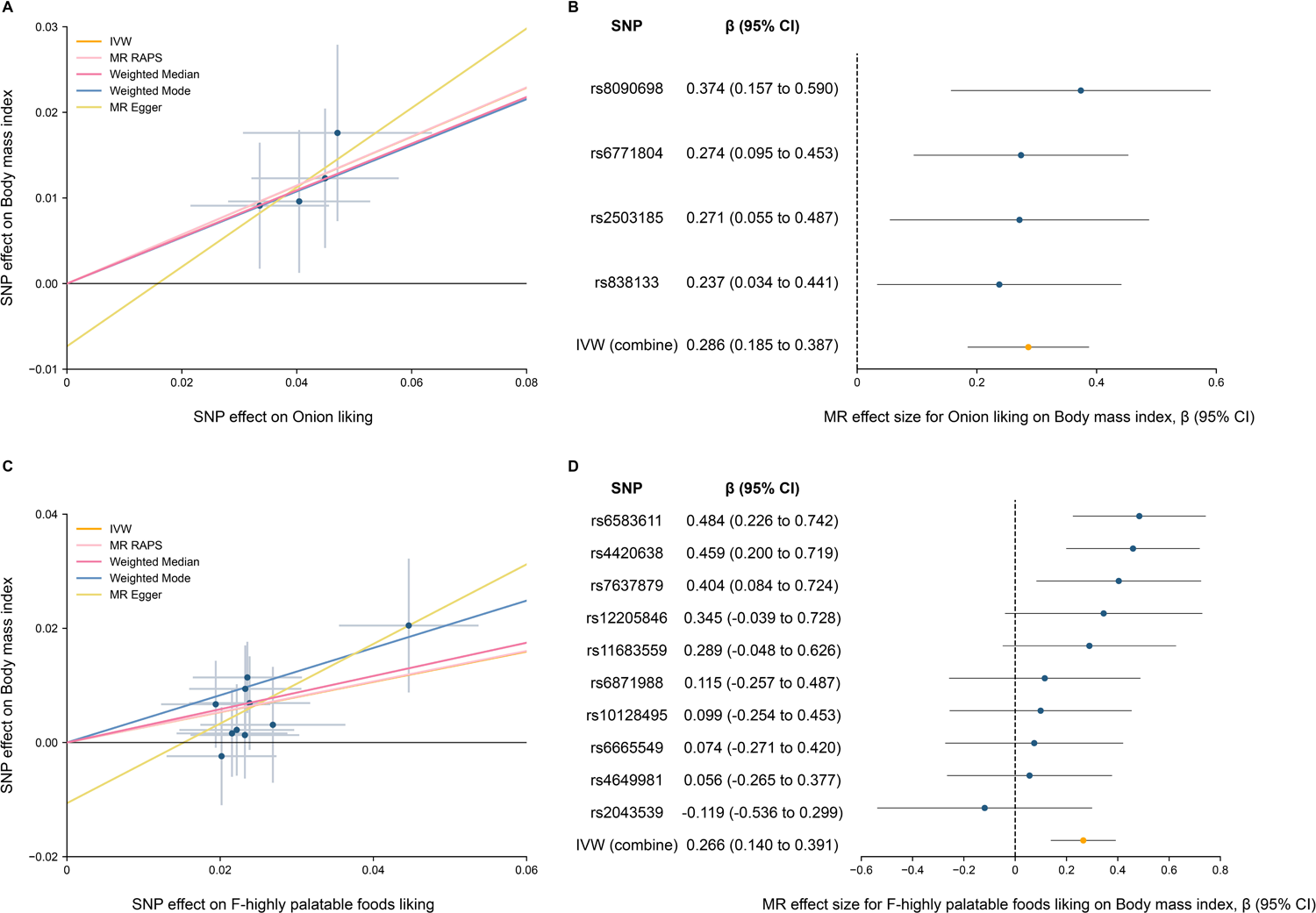
Mendelian randomization (MR) plots for food liking on body mass index (BMI). (A) Scatterplot of single-nucleotide polymorphism (SNP) effects on onion liking and BMI, and the slope of each line corresponding to estimated MR effect per method. (B) Forest plot of individual and combined SNP MR-estimated effects sizes of onion liking on BMI. (C) Scatterplot of SNP effects on highly palatable foods liking and BMI. (D) Forest plot of individual and combined SNP MR-estimated effects sizes of highly palatable foods liking on BMI.

Using genetic liability for obesity phenotypes as the exposures, we conducted MR analyses to investigate the causal effect of obesity-related phenotypes on food liking. Detailed information regarding the SNPs included and excluded for obesity can be found in Supplementary Tables 11-14. Reverse MR analyses revealed a positive association between genetically predicted BMI and onion liking (IVW β, 0.164; 95% CI, 0.085 to 0.243; *P*=4.55×10^−5^), while no significant association was observed with highly palatable foods liking (IVW β, −0.010; 95% CI, −0.056 to 0.035; *P*=0.66) (Supplementary Table 15, Supplementary Figures 2-3). Sensitivity analyses did not identify any pleiotropy (Supplementary Table 16). Leave-one-SNP-out analysis did not detect any influential points, and funnel plots suggested our results were less likely to be influenced by potential biases, with SNPs symmetrically distributed (Supplementary Figures 2-3).

To assess whether specific food preferences might affect obesity in the absence of actual food consumption, we conducted a multivariable MR analysis to estimate the direct effect of identified food preferences on obesity while conditioned on corresponding reported consumption. Unfortunately, the absence of comparable food consumption GWAS for higher-order factors of hierarchical food liking makes multivariable MR analyses impossible. Therefore, we solely evaluated the effect of onion liking on BMI while conditioned on onion intake. The independent instruments are listed in Supplementary Tables 17-18. The effect estimated for onion liking on BMI was consistent with the univariable IVW estimate (univariable IVW β, 0.286; 95% CI, 0.185 to 0.387; *P*=2.80×10^−8^; multivariable IVW β, 0.170; 95% CI, 0.078 to 0.261; *P*=3.41×10^−3^) (Supplementary Table 19).

### Shared loci between identified food liking and obesity

We utilized pleiotropic analysis under the composite null hypothesis (PLACO) ^43^ to identify potential pleiotropic variants associated with the two identified trait pairs. We identified 14 pleiotropic loci for onion liking-BMI and 18 pleiotropic loci for highly palatable foods liking-BMI, including four regions identified in both pairs of traits (*P* < 5×10^-8^) (Figure 4, Supplementary Table 20). Overall, 20 top SNPs in 32 pleiotropic loci exhibited concordant association (Supplementary Table 21), suggesting they were simultaneously positively or negatively associated with specific food liking and BMI. Additionally, further colocalization analysis identified six potential pleiotropic loci with the PP.H4 larger than 0.7 (Supplementary Table 22). Regional plots for each trait pair are depicted in Figure 5. Notably, the 4q24 locus emerged as a pleiotropic locus for both trait pairs, with the same potential shared causal variant, rs13107325, an exonic variant mapped to *SLC39A8*. In contrast, four pleiotropic loci were identified with a PP.H3 larger than 0.7, indicating distinct causal variants between food liking and BMI GWAS.

**Figure 4.**
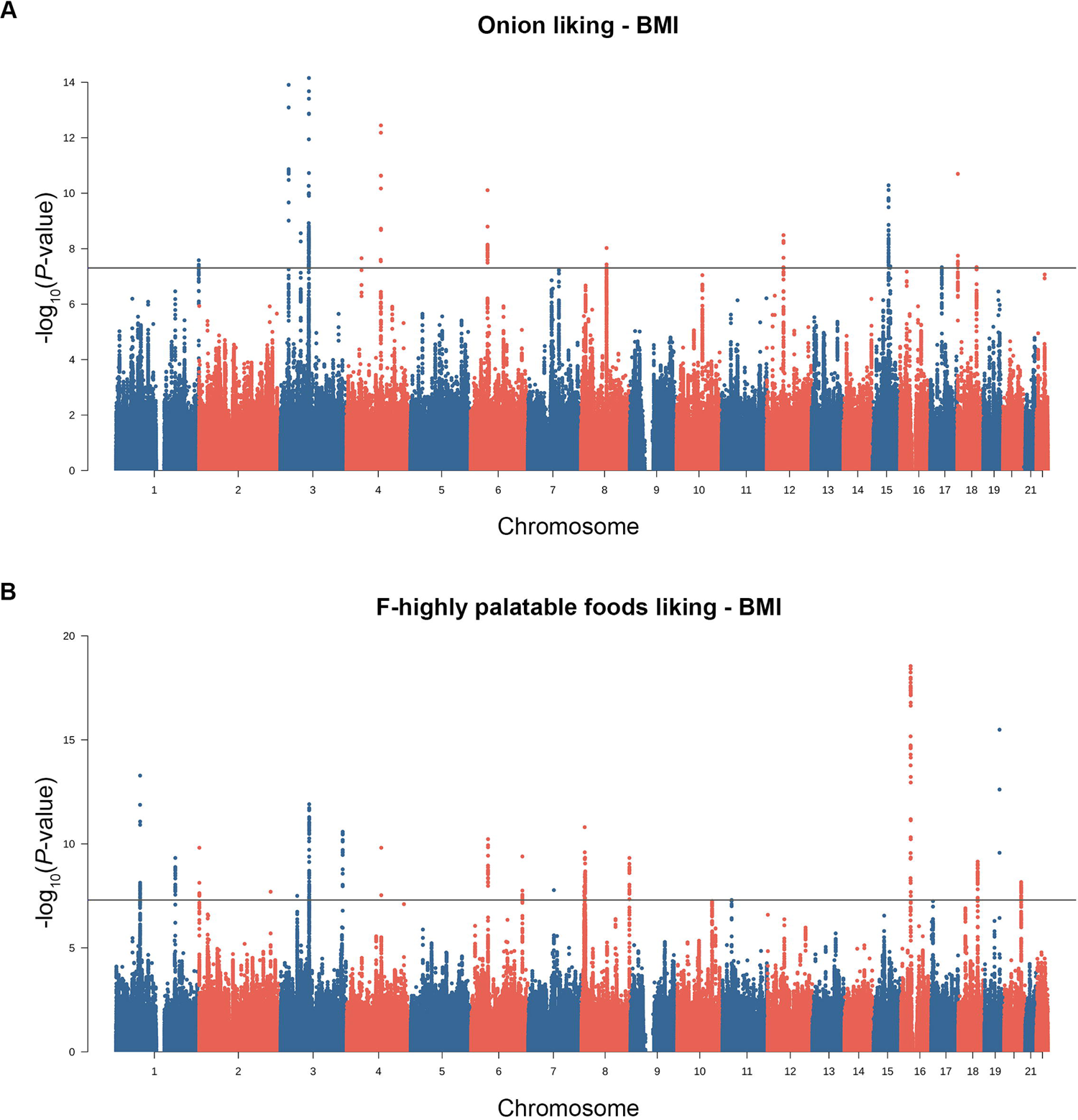
The pleiotropic associations between food liking and body mass index (BMI). (A) Manhattan plots of pleiotropic loci shared between onion liking and BMI. (B) Manhattan plots of pleiotropic loci shared between highly palatable foods liking and BMI. The horizontal gray line indicates the genome-wide significance threshold of 5×10^-8^.

**Figure 5.**
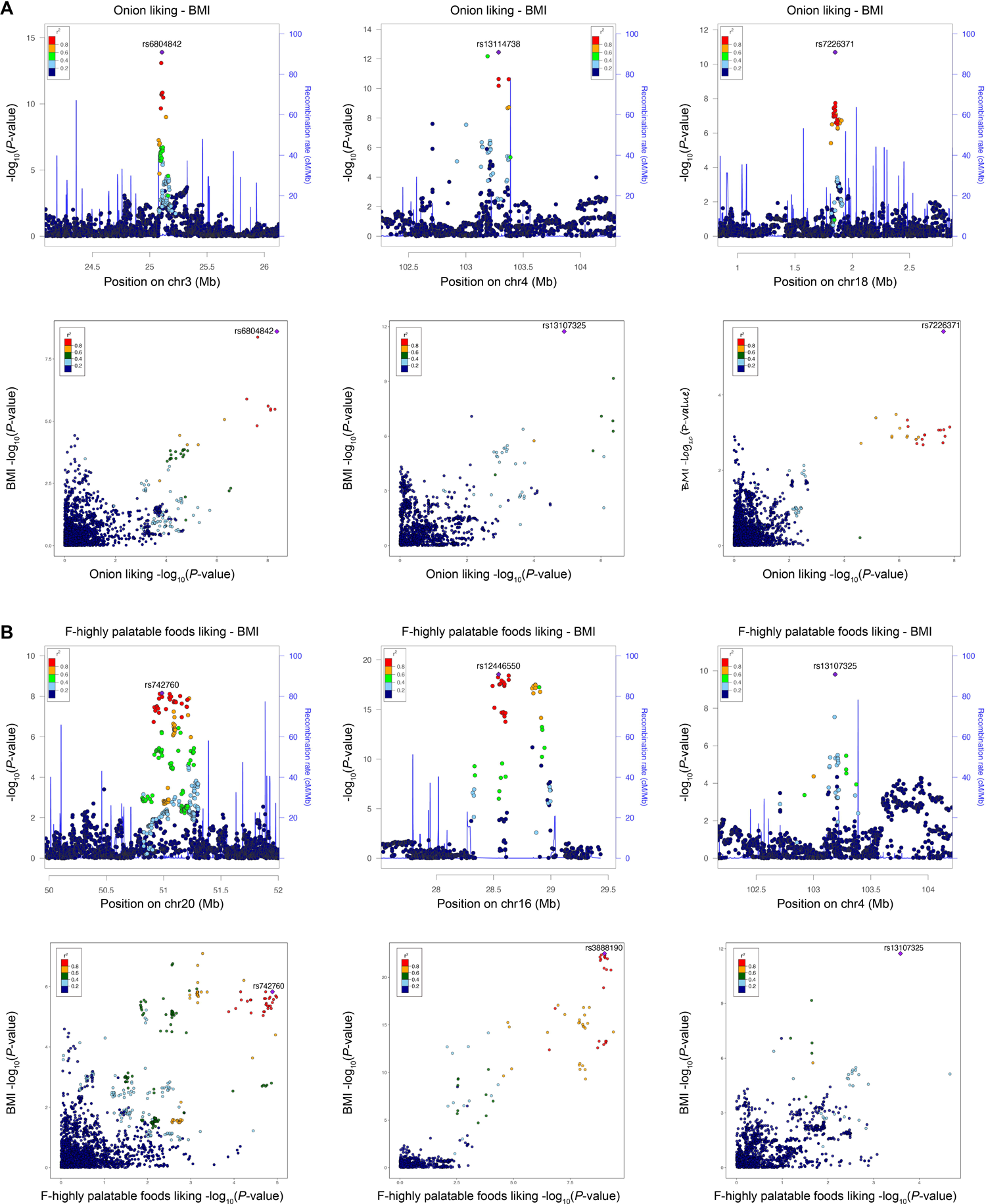
Shared genetic etiology of significantly colocalized loci identified by colocalization analysis. (A) Three significantly colocalized loci for onion liking and BMI. (B) Three significantly colocalized loci for highly palatable foods liking and BMI. The top panels present regional plots visualizing PLACO results for the genomic regions, with lead variants from the pleiotropic analyses labeled. The bottom panels display LocusCompare plots illustrating colocalization between GWAS for food liking and GWAS for BMI. The candidate shared causal variant identified by pairwise colocalization analysis is also labeled. Other SNPs are colored based on their LD r^2^ with the labeled variants.

### Characterization of tissue specificity and prioritization of pleiotropic genes

We employed multi-marker analysis of genomic annotation (MAGMA) ^46^ gene-property analyses to pinpoint tissue specificity, highlighting several brain tissues in both trait pairs (Figure 6A). Subsequently, we applied four methods to prioritize pleiotropic genes, including MAGMA and three methods integrated into FUMA ^44^ (positional, eQTL, and 3D chromatin interaction mapping), utilizing brain eQTL and Hi-C datasets (Figure 6B). A total of 49 genes for onion liking-BMI and 156 genes for highly palatable foods liking-BMI pairs were identified by at least one gene annotation method (Supplementary Tables 23-24). Among these, four genes for onion liking-BMI (*NCKAP5L*, *MAP2K5*, *SKOR1*, *GGNBP2*) and ten genes for highly palatable foods liking-BMI (*RBM6*, *MST1R*, *CAMKV*, *XKR6*, *MSRA*, *TSNARE1*, *SULT1A1*, *NFATC2IP*, *NUPR1*, *ATP2A1*) were highlighted across all four methods. Additionally, the aforementioned *SLC39A8* was also identified for both trait pairs using positional and eQTL mapping.

**Figure 6.**
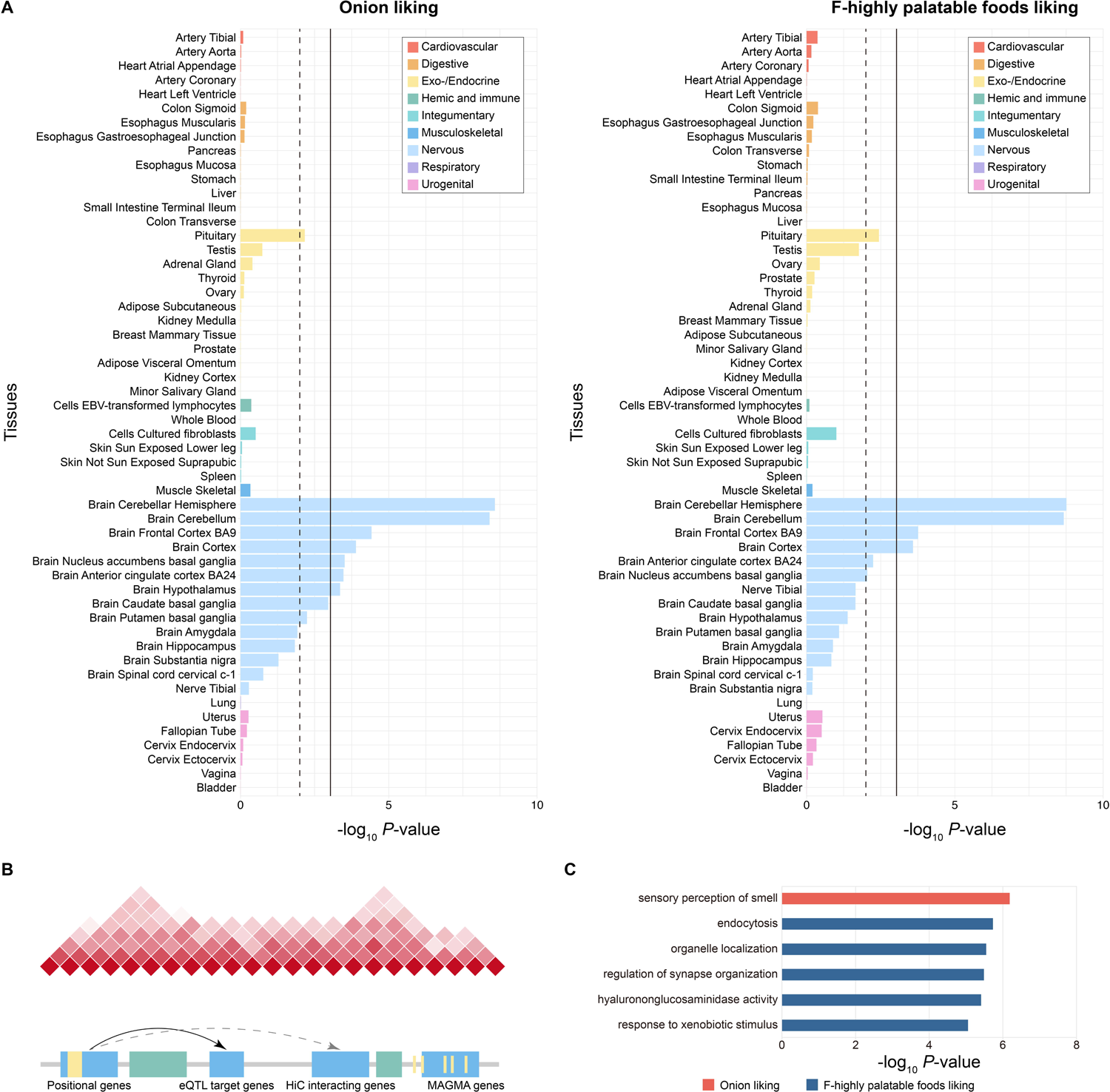
Tissue specificity and gene set enrichment for pleiotropic genes. (A) GTEx tissue-specific enrichment results of the pleiotropic associations. 54 GTEx tissues were classified as nine categories, of which the brain tissues were colored in light blue. The solid line represents the *P*-value threshold after Bonferroni correction (*P* = 0.05/54 = 9.26×10^-4^) and the dotted line represents the nominally significant threshold of *P*-value < 0.05. (B) Four gene prioritization strategies were used for significantly associated loci. Candidate genes were mapped if the pleiotropic loci reside within a protein-coding gene, or eQTL markers of the gene in the brain tissue, or interact with the gene based on brain Hi-C data, or identified utilizing the multi-marker analysis of genomic annotation (MAGMA). (C) Pathway enrichment analysis based on Gene Ontology (GO) terms and Kyoto Encyclopedia of Genes and Genomes (KEGG) pathways among genes implicated in pleiotropic loci.

Gene-set enrichment analyses uncovered six significantly enriched pathway groups (enrichment factor > 2 and adjusted *P* < 0.05) (Figure 6C). For instance, our findings suggested that the involvement of pleiotropic genes associated with onion liking and BMI in sensory perception of smell (*P* = 6.52×10^-7^) might explain how onion liking affects obesity. Pleiotropic genes for highly palatable foods liking and BMI were enriched in five pathways, including endocytosis (*P* = 1.88×10^-6^) and organelle localization (*P* = 2.83×10^-6^).

## Discussion

Addressing the ongoing increase in obesity prevalence demands multifaceted and effective responses for both prevention and treatment. Genetic correlations have been suggested between preference for low-caloric and acquired food and lower obesity indices. In contrast, the preference for highly palatable foods has the opposite pattern ^13^, but the causal relationship has not been investigated. In this study, we attempted to disentangle relationships between food liking and obesity risk using genome-wide cross-trait analysis. We observed evidence indicating that liking highly palatable foods and onions would causally increase BMI and the effect of onion liking on BMI independent of actual onion intake. Further comprehensive analyses highlighted potential pleiotropic genetic loci, shared causal variants, pleiotropic genes, and biological pathways, supported shared genetic etiology underlying liking for onions and highly palatable foods and BMI.

Dietary preferences are multifaceted and influenced by various factors such as genetic inheritance and cultural and social aspects ^13,49^, and the food sensory properties are perceived through a combination of taste, texture, and olfaction. Highly palatable foods capture our attention and can lead to at least transient increases in consumption because of their high hedonic value ^50^. Evolutionarily, humans had to survive undernutrition periods. Thus, selection pressure most likely contributed to a genotype that favors highly palatable energy-dense foods ^51^. However, the adoption of relatively sedentary lifestyles combined with the availability of energy-dense, highly palatable foods represents a nutritional shift that has become one of the most significant risk factors for obesity ^4^. Functional magnetic resonance imaging (fMRI) studies have shown that obese individuals often exhibit heightened activation in brain regions that encode food sensory and hedonic aspects when exposed to palatable food images or anticipated receipt of palatable foods ^52,53^. These changes in the neural reward homeostasis might lead to the development of addictive-like behavior, leading to compulsive-like feeding behavior and subsequent overconsumption and weight gain ^8^. Understanding how highly palatable food liking influences eating behavior and contributes to obesity risk is crucial for developing effective interventions to address the obesity epidemic.

We identified the causal effect of liking highly palatable foods and onions on increased BMI, and a wide range of studies found that dietary preferences are one of the most important drivers of food consumption ^54^. However, through multivariable MR, we found that the effect of onion liking on BMI remains significant even after conditioning on onion intake, indicating a certain degree of independence in the influence of dietary preferences. The onion liking falls into the category of savory food liking and contributes to both highly palatable food liking and acquired food liking ^13^. If food preference indeed has an independent impact on obesity, it would not be surprising for intervention plans focused on changing diet and physical activity to lose effectiveness in the long term ^2^. Nowadays, alternatives with the sensory properties of highly palatable foods, such as non-nutritive sweeteners, have been used in the food industry to reduce the global obesity problem. However, such replacement has been suggested to promote the accumulation of adipose tissue and result in weight gain ^55^. One plausible explanation is the potential decrease in satiety and the subsequent increase in preference for palatable foods, which could impact overall dietary patterns ^56^. Considering the possible independent impact of food liking and food liking is not stable within individuals ^6^, obesogenic marketing to promote foods high in sugar and fat should be limited. Such advertisements may increase preference for energy-dense foods and beverages ^57^.

Pleiotropic variants between food liking and obesity exhibited extensive distribution, with numerous loci highlighted between two identified trait pairs. Several previously identified loci associated with food liking were illustrated to be potential pleiotropic loci shared with BMI. For example, *OR2M3* (1q44) is a highly specific and narrowly tuned human odorant receptor crucial for detecting the key odorant in onions ^58^, yet no studies to date have reported the associations of *OR2M3* with obesity phenotypes. Pleiotropic gene prioritization also highlighted several olfactory receptors and enriched the pathway of sensory perception of smell, indicating the critical role of olfactory behavior in food preference and obesity. In the meantime, we also identified several loci previously linked to obesity, which were identified as potential pleiotropic loci shared with food liking. For instance, *SLC39A8* (4q24) was suggested to be shared between onion liking and BMI as well as highly palatable food liking and BMI. The potential shared causal nonsynonymous variant in *SLC39A8* (rs13107325) is among the most pleiotropic variants in the human genome ^59,60^. It has been associated with various diseases including obesity, schizophrenia, Parkinson’s disease, and others. Moreover, the *SLC39A8* gene was reported as a shared gene between obesity and the development of subcortical brain structures ^61^, consistent with our tissue specificity results and the neural control of essential behaviors. For example, opioidergic circuits in regions like the nucleus accumbens and the ventral pallidum encode hedonic liking ^4^.

It is crucial to recognize the limitations of our study when interpreting the results. Our study utilized the largest GWAS summary statistics for onion intake to perform multivariable MR. However, the genetic factors can only explain a small proportion of the variance in food composition, and we employed a relaxed *P*-value threshold to select genetic instruments due to the limited number of variants, which may introduce weak instrument bias. Nevertheless, our rigorous analysis included testing instrument strength, revealing that all instruments had *F* statistics exceeding 10, a conventional cutoff for robust instruments ^20^. Moreover, the absence of comparable food consumption GWAS for higher-order factors of hierarchical food liking makes multivariable MR analyses impossible for highly palatable food liking. Second, our study was confined to individuals of European ancestry to avoid potential confounding due to ancestral heterogeneity, potentially limiting the generalizability of our findings to other ancestral populations. Third, SNPs linked to food liking were primarily identified in the UK Biobank study, focusing on individuals aged 40 to 69, whereas the obesity GWAS samples encompassed a broader age range. Younger individuals’ dietary preferences might be influenced by other variants with varying associations with obesity, though such GWAS data is currently unavailable.

In conclusion, we conducted a cross-trait analysis to explore the relationship between 187 food-liking and obesity. We provide compelling evidence suggesting that onion liking and highly palatable food liking are linked to increased BMI, with onion liking exerting an independent effect on BMI after conditioning on actual onion intake. Furthermore, our comprehensive analyses highlighted a shared genetic etiology underlying food liking and BMI. These findings not only provide a better understanding of the association between food preferences and obesity but also contribute to the formulation of policies.

## Data Availability

All data produced in the present study are available upon reasonable request to the authors.

## Acknowledgements

This study is supported by the National Natural Science Foundation of China (82101601, 82071190, 82371438), the Affiliated Hospital of Guangdong Medical University High level Talent Research Launch Project (GCC2022001), the Innovative Strong School Project of Guangdong Medical University (4SG21230G), and the Open Fund of Guangdong Key Laboratory of Age-Related Cardiac and Cerebral Diseases (2023).

## Author contributions

S.Y. conceived this study, participated in the data collection, and wrote the paper. S.Y., and H.W. performed the data analyses. S.Y., H.W., P.B., L.Q., and J.-Z.H. prepared the tables and figures. Y.W., S.-F.F., Y.-J.C., J.G., X.K., W.S., F.-B.M., Q.H.L., S.-S.D. and Y.G. critically revised the content. D.-L.Z. and L.-L.C. supervised the study. All authors contributed to editing the paper.

