## Supplemental Figures for "A genome-wide cross-trait analysis identifies causal relationship and shared loci of food preference with obesity"

Yao *et al.*

**Supplementary Figure 1. Sensitivity analyses for the Mendelian randomization (MR) analyses of food liking on body mass index (BMI).**

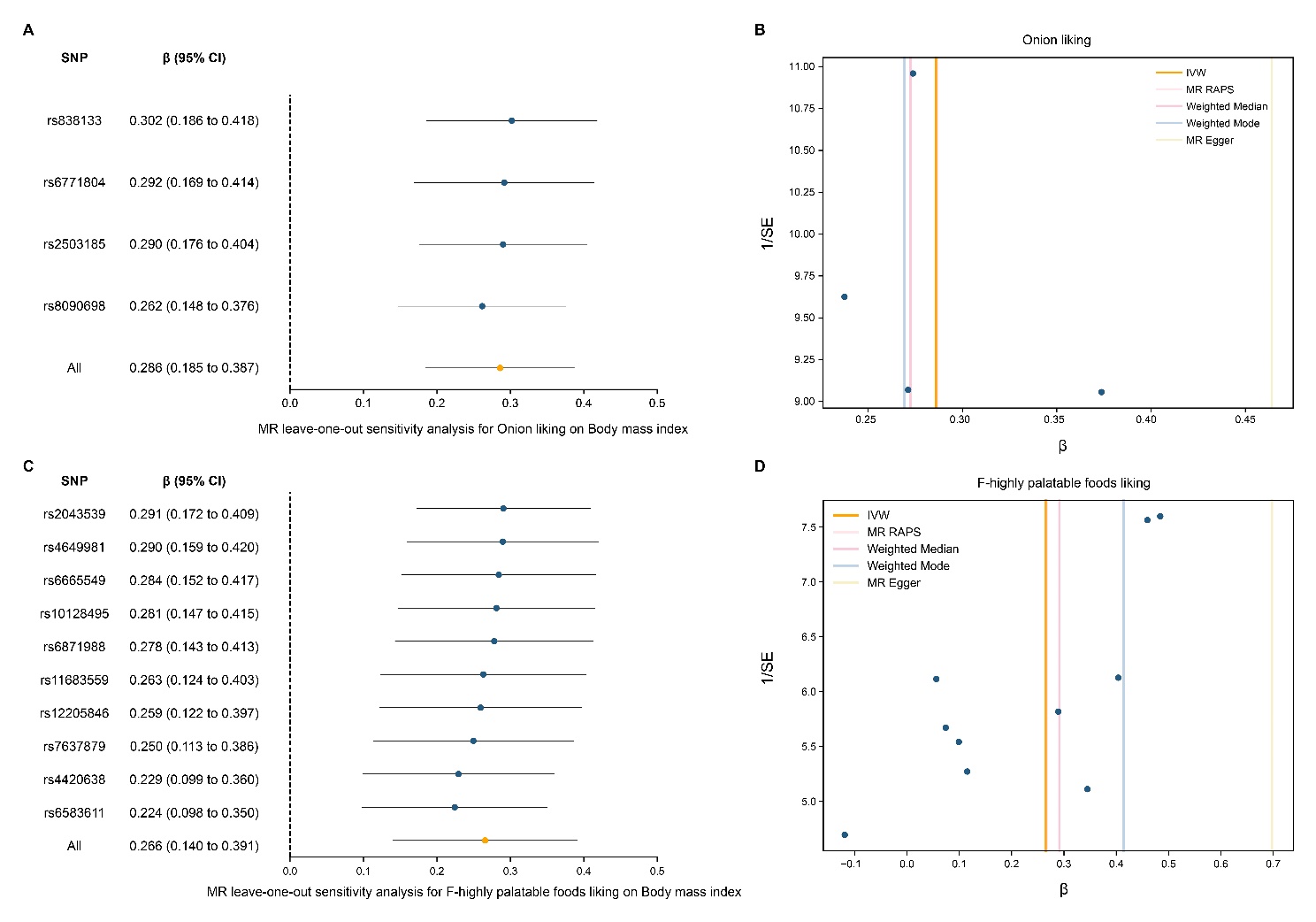

(A) Forest plot of leave-one-out sensitivity analyses to show the influence of individual single-nucleotide polymorphism (SNP) of onion liking on BMI. (B) Funnel plot to visualize overall heterogeneity of MR estimates of onion liking on BMI. (C) Forest plot of leave-one-out sensitivity analyses to show the influence of individual SNP of highly palatable foods liking on BMI. (D) Funnel plot to visualize overall heterogeneity of MR estimates of highly palatable foods liking on BMI.

**Supplementary Figure 2. Mendelian randomization (MR) plots for the relationship of body mass index (BMI) on onion liking.**

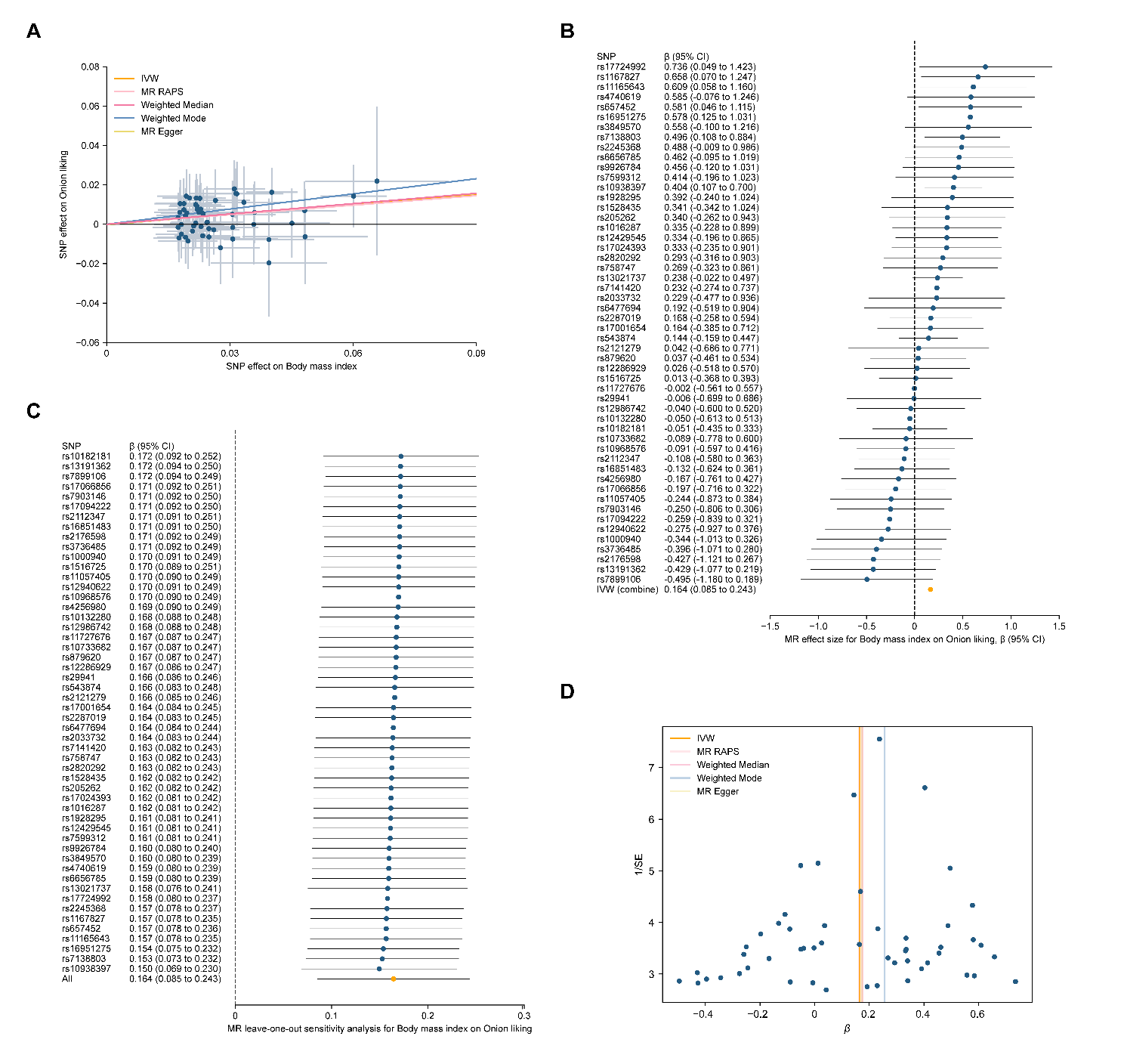

(A) Scatterplot of single-nucleotide polymorphism (SNP) effects on BMI and onion liking, and the slope of each line corresponding to estimated MR effect per method. (B) Forest plot of individual and combined SNP MR-estimated effects sizes. (C) Forest plot of leave-one-out sensitivity analyses to show the influence of individual SNP on the result. (D) Funnel plot to visualize overall heterogeneity of MR estimates.

**Supplementary Figure 3. Mendelian randomization (MR) plots for the relationship of body mass index (BMI) on highly palatable foods liking.**

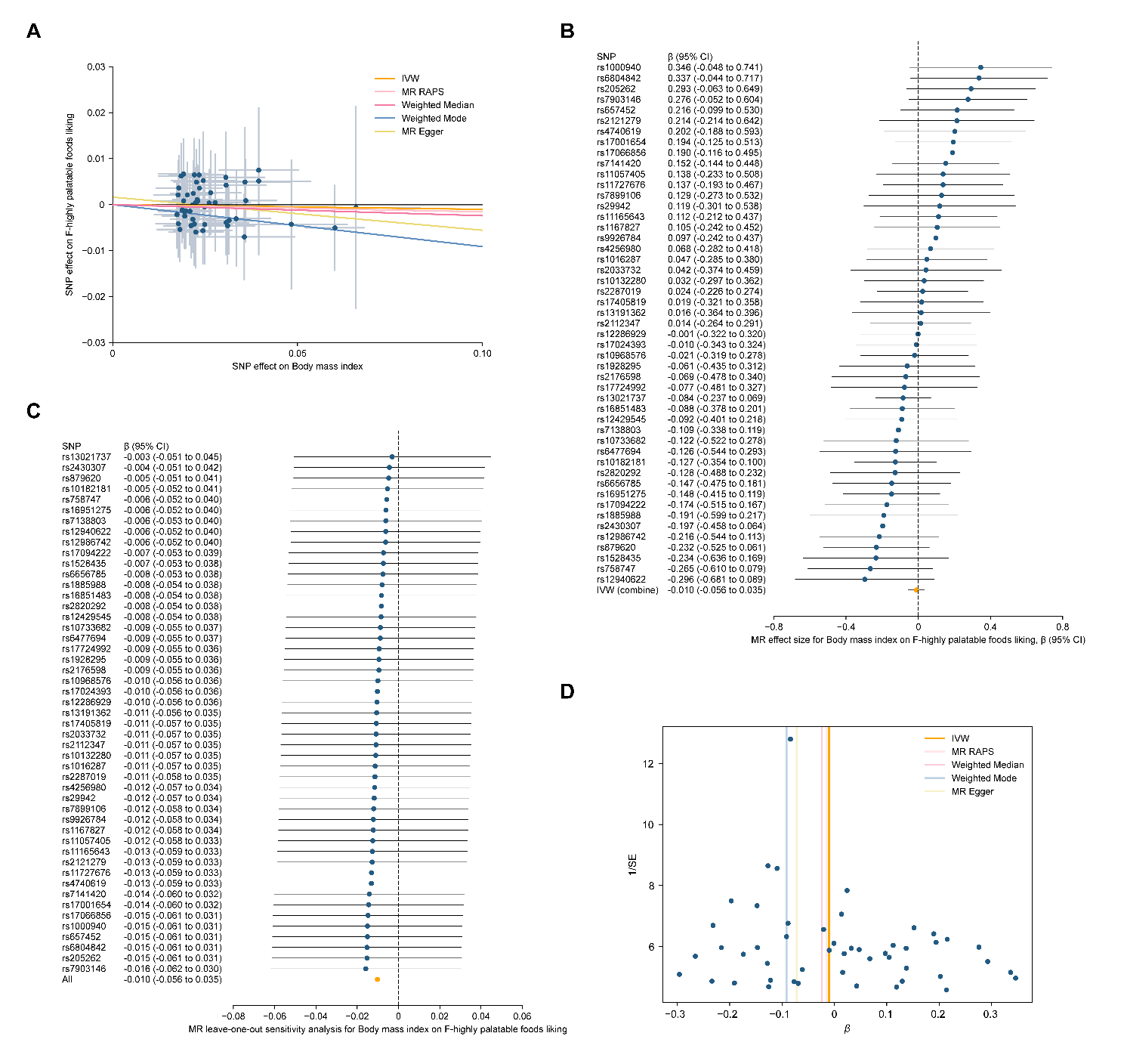

(A) Scatterplot of single-nucleotide polymorphism (SNP) effects on BMI and highly palatable foods liking, and the slope of each line corresponding to estimated MR effect per method. (B) Forest plot of individual and combined SNP MR-estimated effects sizes. (C) Forest plot of leave-one-out sensitivity analyses to show the influence of individual SNP on the result. (D) Funnel plot to visualize overall heterogeneity of MR estimates.
